# Psychometric validation of an Arabic version of the WHO-5 wellbeing scale among Lebanese Adolescents

**DOI:** 10.1101/2025.01.05.25320021

**Authors:** Rita Doumit, Souheil Hallit, Maria-Jose Sanchez-Ruiz, Myriam El Khoury-Malhame

## Abstract

**Introduction:** Wellbeing in adolescence is frequently associated with positive developmental outcomes. The WHO-5 is widely recognized for its brevity, clarity, robust reliability, and cultural validity. In this study, we aimed to assess the psychometric properties of an Arabic version of the WHO-5 scale among Lebanese adolescents.

**Methods:** This cross-sectional study involved 681 Lebanese school students aged 15-18 years. Participants were assessed using WHO-5 (wellbeing), PHQ-9 (depression), and GAD-7 (anxiety) Arabic questionnaires at two-time points, 3 months apart.

**Results:** We found that the 5 items of the WHO-5 converged into a single factor. Composite reliability of scores was adequate in the total sample (ω = .83 / α = .83). The convergent validity for this model was satisfactory. We were able to show the invariance across gender at the configural, metric, and scalar levels, with males showing a higher level of wellbeing compared to females. The pre-posttest assessment for the WHO-5 scale was conducted on 358 participants; the intraclass correlation coefficient was adequate = 0.78 [95% CI .73; .82]. Our analyses also show that wellbeing was negatively correlated with depression (r = −.54; p < .001) and anxiety (r = −.52; p < .001).

**Conclusion:** The Arabic WHO-5 among Lebanese adolescents displayed highly satisfactory psychometric properties, which are evidence of its validity. It could be used to better track positive mental health in this vulnerable age group and could highlight efficiency in interventions aiming to promote wellbeing in adolescents. It could also potentially identify at risk individuals.

## Introduction

Wellbeing is a multifaceted construct mostly linked to positive mental health, emotional stability, and psychological wellness (1). The World Health Organization emphasizes its definition as a “state of complete physical, mental and social wellbeing and not merely the absence of disease or infirmity” (2). In the absence of unified definitions for wellbeing and with the lack of clear biomarkers, various scales have been validated to measure this construct. The most widely used versions include, yet are not restricted to, the World Health Organization Wellbeing Index (WHO-5) (3), General Wellbeing (4), the Wellbeing Numerical Rating Scales (WB-NRSs) (5) and Psychological Wellbeing Scale (PWB) measuring 6 aspects of wellbeing (6).

Other scales are known to assess conceptually similar constructs such as life satisfaction, positive affect, optimism, and happiness (7). Among the validated wellbeing-related scales, the WHO-5 stands out for its brevity and ease of use, with only 5 items as opposed for instance to the PWB (42 items) and even its shorter version (18 items). It has strong psychometric properties, minimal overlap with scales measuring mental distress, and has also been translated into over 30 languages. Its adoption has been advocated by the WHO-led recommendations, with the aim of facilitating cross-cultural comparisons of the prevalence of mental wellness and distress (5). Although this scale has been used in various nations and clinical settings, it has mostly been validated in high-income, western countries and only a few studies have targeted the Middle East (8) or particularly adolescents (9).

Adolescence is a critical developmental stage marked by blatant physical changes, driven by hormonal fluctuation, and often accompanied by emotional lability and interpersonal conflicts (10). As young adolescents learn to navigate those changes, they become more susceptible to psychological distress and decreased wellbeing (11; 12). In this age group, there is a wealth of evidence linking wellbeing with various developmental benefits, including improved academic performance, reduced risks of mental health problems, stronger social ties, and increased resilience (13; 14). A few studies have investigated some of these benefits in Lebanese youth, especially those related to positive educational outcomes (15).

Measuring wellbeing using WHO-5 in the adolescent population have been conducted in Ghana (16), Europe (17), and ethnic minorities in Norway and Sweden (18). However, despite its widespread use, there is a surprising gap in validation studies within the, Arab region, which is known to host one of the most youthful age structures globally (45, 14). From a cultural perspective, adolescents’ distress is particularly prominent in societies marked by collective traumas, often indicating an increased risk of psychiatric disorders as they transition into young adulthood (19). Lebanon, as a small Middle Eastern country, bears the weight of accumulated national adversities stemming from decades of civil war, including severe socio-political unrest, the most devastating economic collapse of its modern history, and a cataclysmic explosion of the Beirut port. These challenges unfold against the backdrop of ongoing adjustments to the COVID-19 pandemic and the influx of nearly 1.5 million Syrian refugees (20). Taken together with failing governmental institutions and a lack of mental health public policies, these crises have left the country with staggering psychological distress levels (21; 22) and plummeting levels of wellbeing (23). Recent studies particularly point to alarming levels of anxiety and depression in Lebanese adolescents reaching more than 60% for anxiety and 35% for depression after the Beirut blast (19), while others have focused on protective factors of wellbeing among Lebanese youth during the COVID-19 pandemic, such as trait Emotional Intelligence and meaning-centered coping (24).

In line with global concerns around adolescents’ deteriorating mental health (25) and growing evidence of worsening their distress and isolation (24), determining levels of wellbeing in a vulnerable age group becomes of utmost importance, notably when taken against a backdrop of sociopolitical, healthcare, and financial instabilities. Given the dynamic nature of subjective wellbeing, it would allow the design of targeted interventions aimed at increasing its levels of wellbeing to bolster psychological distress. This study thus aims at validating a culturally-adapted Arabic version of the WHO-5, a brief reliable instrument for wellbeing in adolescents. It contributes to assessing and subsequently promoting wellbeing in the future life trajectories of the Lebanese population and in the Arab-speaking context in general.

## Methods

### Participants

The sample consisted of 701 high school students aged between 15 and 18 (mean age 16.31 ± 1.18), who were recruited from four public schools in Lebanon. Some participants were tested twice for the same scales, within 3 months, to assess test-retest validity and reliability. Among those participants, 681 students, years had complete data and were included in the final sample with 56 % females. At retest, 358 participants were included.

### Procedure

The study was part of a larger project; Yes to Emotions in Youth (YEY): An Emotional Intelligence Training for Vulnerable Youth in Lebanon funded by Grand Challenges Canada – Global Mental Health. It received administrative approval from the Lebanese Ministry of Education and Higher Education (MEHE) and was approved by the Institutional Review Board (LAU IRB; reference number: LAU.SAS.MJ1.28/Oct/2021), in accordance to the Declaration of Helsinki. All participants and their parents signed the informed consent before enrolling in the study. Students were asked about their age and gender then filled out the validated publicly available Arabic version of the standardized scales for wellbeing (WHO-5), depression (PHQ-9), and anxiety (GAD-7), in that order.

### Questionnaires

#### WHO-5

(World Health Organization Wellbeing Index; (26)). It is a 5-item questionnaire measuring general emotional wellbeing in the past two weeks. Participants answer Likert-scale questions such as “I have felt cheerful and in good spirits” ranging from 0 “Not at all” to 5 “All the time”. Higher scores indicate higher states of wellbeing and are transformed to a 100-point basis. It has been shown to hold a reliability greater than (α = .8) and has been validated in an Arabic population (8).

#### GAD-7

(The Generalized Anxiety Disorder; (27)). It is a self-reported questionnaire designed to assess anxiety during the previous 2 weeks. It is based on 7 items “e.g., Feeling nervous, anxious, or on edge”, which are responded to on a 4-point Likert scale ranging from 0 “Not at all” to 3 “Nearly every day”. Higher scores indicate higher severity of symptoms. It has satisfactory internal consistency (α from .79 – .91;) and has been validated among an Arabic Lebanese population (28).

#### PHQ-9

(Patient health questionnaire-9; (29)). It is a 9-item questionnaire measuring the frequency of depressive symptoms over the past two weeks, with items such as having “I have little interest or pleasure in doing things” evaluated on a Likert-scale ranging from 0 “Not at all” to 3 “Nearly every day”. Higher scores indicate higher severity of symptoms. It has strong reliability with (α = .82), and was validated in the Lebanese population (28).

### Analytic Strategy

#### Confirmatory factor analysis

There were no missing responses in the dataset. We used data from the total sample to conduct a CFA using the SPSS AMOS v.29 software. We aimed to enroll a minimum of 100 adolescents following the recommendations of Mundfrom et al. of 3 to 20 times the number of the scale’s variables (30). Parameter estimates were obtained using the maximum likelihood method. Multiple fit indices were calculated: Steiger-Lind root mean square error of approximation (RMSEA), standardized root mean square residual (SRMR), Tucker-Lewis Index (TLI), and Comparative Fit Index (CFI). Values ≤ .08 for RMSEA, ≤for SRMR, and ≥ .90 for CFI and TLI indicate good fit of the model to the data (31). Additionally, values of the average variance extracted (AVE) ≥ .50 indicated evidence of convergent validity (32). Multivariate normality was not verified at first (Bollen-Stine bootstrap p= .002); therefore, we performed non-parametric bootstrapping procedure.

#### Gender invariance

To examine gender invariance of WHO-5 scores, we conducted multi-group CFA (33) using the total sample. Measurement invariance was assessed at the configural, metric, and scalar levels (34). We accepted ΔCFI ≤ .010 and ΔRMSEA ≤ .015 or ΔSRMR ≤ .010 as evidence of invariance (35).

#### Further analyses

Composite reliability was assessed using McDonald’s ω and Cronbach’s α, with values greater than .70 reflecting adequate composite reliability (36). Normality was verified since the skewness and kurtosis values for each item of the scale varied between −1 and +1 (37). Pearson test was used to correlate the WHO-5 scores with the other scales in the survey. Student t-test was used to compare two means.

## Results

### Participants

Seven hundred-eight adolescents participated in this study. The mean and standard deviation of the scores were as follows: WHO-5 wellbeing (pre: 41.94 ± 22.62, post: 39.68 ± 21.24), PHQ-9 depression (11.13 ± 6.80), and GAD-7 anxiety (9.61 ± 5.48). The description of the categorization of participants according to their PHQ-9 and GAD-7 scores is summarized in Table 1.

**Table 1.** Categorization of participants according to their depression and anxiety scores.

|  |  |
| --- | --- |
| Depression severity |  |
| No depression (PHQ-9 scores 0-4) | 137 (19.4%) |
| Mild (PHQ-9 scores 5-9) | 196 (27.7%) |
| Moderate (PHQ-9 scores 10-14) | 146 (20.7%) |
| Moderately severe (PHQ-9 scores 15-19) | 132 (18.7%) |
| Severe (PHQ-9 scores 20-27) | 96 (13.6%) |
| Anxiety severity |  |
| No anxiety (GAD-7 scores 0-4) | 148 (20.9%) |
| Mild (GAD-7 scores 5-9) | 229 (32.3%) |
| Moderate (GAD-7 scores 10-14) | 178 (25.1%) |
| Severe (GAD-7 scores $\geq 15$) | 153 (21.6%) |

### Confirmatory Factor Analysis of the WHO-5 scale

CFA indicated that fit of the one-factor model of the WHO-5 scale was acceptable: RMSEA = .095 (90% CI .068, .125), SRMR = .028, CFI = .974, TLI = .949. The standardized estimates of factor loadings were all adequate (Figure 1). Composite reliability of scores was adequate in the total sample (ω = .83 / α = .83). The convergent validity for this model was satisfactory, as AVE = .51.

**Figure 1.**
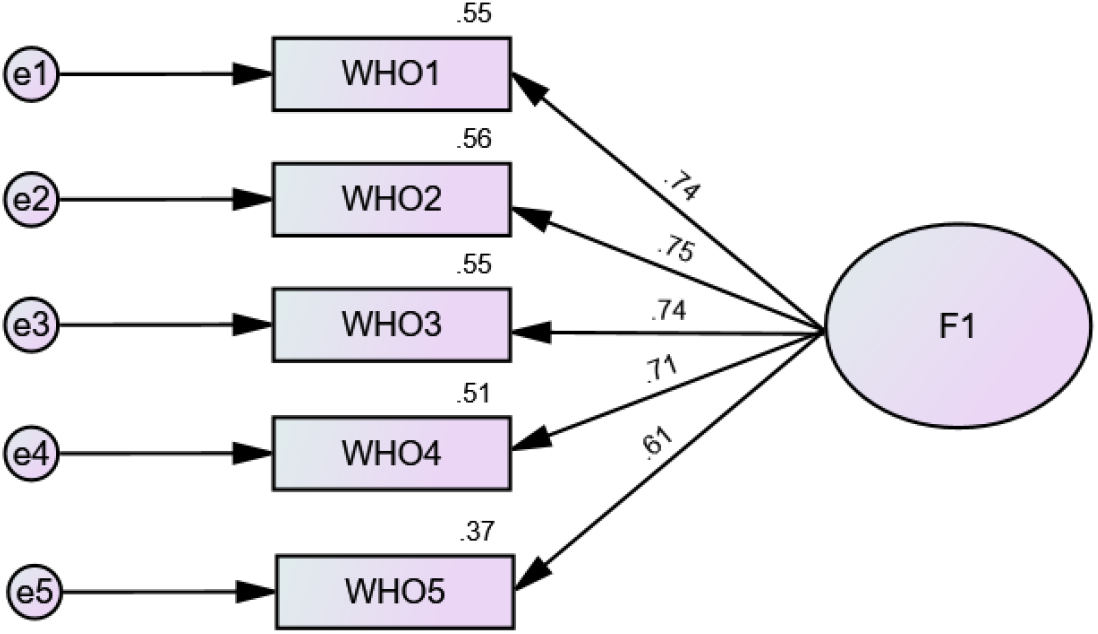
Standardized loading factors of the World Health Organization Wellbeing 5 items Scale items in Arabic.

### Gender Invariance

We were able to show the invariance across gender at the configural, metric, and scalar levels (Table 2). Males showed a higher level of wellbeing compared to females (11.43 ± 5.96 vs 9.81± 5.30; *t*(679) = 3.68; *p* < .001).

**Table 2.** Measurement Invariance of the World Health Organization Wellbeing 5-item Scale across gender in the total sample.

| Model | CFI | RMSEA | SRMR | Model Comparison | ΔCFI | ΔRMSEA | ΔSRMR |
| --- | --- | --- | --- | --- | --- | --- | --- |
| Configural | .970 | .072 | .030 |  |  |  |  |
| Metric | .972 | .059 | .031 | Configural vs metric | .002 | .013 | .001 |
| Scalar | .967 | .056 | .031 | Metric vs scalar | .005 | .003 | <.001 |

*Note.* CFI = Comparative fit index; RMSEA = Steiger-Lind root mean square error of approximation; SRMR = Standardized root mean square residual.

### Interclass Correlation

The pre-posttest for the WHO-5 scale was done on 358 participants. The intraclass correlation coefficient was adequate = 0.78 [95% CI .73; .82]

### Concurrent validity

Higher depression (r = −.54; p < .001) and anxiety (r = −.52; p < .001) were significantly associated with lower wellbeing.

## Discussion

This study investigated the psychometric properties of the Arabic version of the WHO-5 scale in a group of Arabic-speaking Lebanese high-school students. Findings support a single-factor model, with adequate reliability (McDonald’s ω = .93), as well as good convergent and divergent validity. Measurement invariance across gender groups was also evidenced at the configural, metric, and scalar levels. Consequently, the WHO-5 could be exploited as a reliable and valid tool, time-effective and easy to use, with cultural sensitivity to evaluate Arab adolescents’ overall wellbeing. Given that this scale has been mostly administered in adults in Western countries, investigating its validity among Lebanese adolescents further endorses its cross-cultural potential and its suitability for measurements of wellbeing in Arabic-speaking adolescents. This could also be particularly relevant for other countries sharing some of the overall settings (20), including vulnerable populations, low-income countries, and those plagued by atrocities of political unrest, ongoing wars, and/or severe monetary depreciation.

The WHO-5 scores in our sample of 681 school students aged 15-18 years were found to be lower compared to means documented in an international study among European adolescents with comparable age range and gender distribution (17). Those averages were nonetheless comparable to those of young adults in Lebanon and in other low-middle-income Arab countries such as Morocco, Kuwait, and Jordan (8). Taken together with values obtained on anxiety (GAD-7) and depression scales (PHQ-9), these results indicate elevated levels of mental distress among Lebanese-educated adolescents, with low levels of wellbeing, and heightened psychological distress in participants; reporting moderate to severe symptoms of anxiety (53%) and depression (47%).

Our findings are in line with recent national prevalence rates of mental distress in adolescents after the Beirut port explosion (19). This study, conducted a few years post-blast and post-pandemic, illustrates the chronicity and lasting impact of accumulating adversities on adolescents. Although alarming, those findings might be expected in the backdrop of a prolonged collective challenges and traumas altering the quality of life, of the Lebanese people in general and on the most vulnerable youngsters in particular (38). These crises are aggravated by the absence of governmental policies addressing mental health and the scarcity of school-based mental health interventions promoting wellbeing and interpersonal coping skills. As such, results should inform altogether clinicians, researchers, and policymakers to implement culturally sensitive strategies aiming at addressing wellbeing as well as managing anxious and depressive symptomatology among Lebanese adolescents to prevent potential worsening of mental and physical health.

Per se, the psychometric properties of the WHO-5 construct validity were examined using CFA, an approach systematically supported by validation researchers as an essential step in scale validation (39). Our analyses showed the fit of a one-factor model, replicating the single-factor structure of the original scale version (40). Similarly, subsequently, psychometric studies across different countries (5) and different age groups including Arab adults and adolescents (16; 18) have consistently supported the unidimensional structure. This could infer that the assessment of adolescents’ wellbeing in Lebanon could be accounted for by a single underlying factor, comparably to age-matched participants from other countries. It mostly supports the applicability of the WHO-5 scale within the Lebanese setting as a unidimensional measure of wellbeing and its recommendation for use in Arab adolescents globally. Future studies could additionally investigate whether current findings within the general population could be replicated in clinical populations of adolescents, whereby a single latent trait would also help evaluate their wellbeing.

Moreover, the unidimensional factor structure of this Arabic version of the WHO-5 was identical for both male and female high schoolers at the three levels of invariance (Configural, Metric, and Scalar). We found no significant gender differences in scores, and as such our results are in support of indiscriminately using the WHO-5 across genders, aligning with findings from previous studies (9). The WHO-5 items also appeared to be internally consistent, and quite similar to values obtained in a recent validation of the same Arabic version of WHO-5 in a sample of non-clinical Arab adults across six Arab countries including Lebanon (8). One supplementary distinctive feature of this study was its evaluation of test-retest interclass correlation of wellbeing measures since more than half of the original sample of adolescents were retested at 3 months post-initial assessment. Results point to statistically adequate values and further evidence of the reliable psychometric properties of WHO-5 in Arab adolescents.

Lastly, the WHO-5 scores showed a strong and negative correlation with depression and anxiety scores. These findings also concur with prior research comparing the correlations between the WHO-5 with various measures of mental distress and related problems worldwide and more precisely in Middle Eastern and neighboring countries (41; 42; 8).

### Implications, Limitations and Recommendations for Future Research

Our results suggest that the WHO-5, Arabic version of the WHO-5 can be a valuable tool for researchers and clinicians aiming to assess wellbeing among Arab-speaking adolescents. This scale offers a quick, convenient, and age-appropriate means of assessing wellbeing, with the distinct advantage of being culturally sensitive and easy to understand. In addition, recent studies have highlighted that the WHO-5 could also serve as a proxy measure for clinical depression in the general population (43; 44). As such, psychologists, public health nurses, and educators could use it to assess wellbeing, and, by the same token, depressive symptoms among high schoolers, particularly those living in vulnerable environments.

This study supports the use of the WHO-5 Arab version in young adolescents in a Middle Eastern country. Although psychometric properties indicate valid and reliable usage of the scale in a culturally sensitive manner, results should be generalized with care to other Arabic-speaking adolescents, especially since the sample included in the study was educated urban youth. Further research should as such target larger sample sizes and non-schooled youth. Additional limitation stems from relying solely on self-report measures that may introduce known biases such as social desirability, memory inaccuracies, and potential misinterpretations of items. Nonetheless, the test-retest solid reliability, gender invariance, and religious as well as regional diversity representation within the targeted public schools in Lebanon could mitigate such biases. Future studies should further investigate the potential influence of other demographic factors on wellbeing, incorporate physiological markers, and potentially study clinical samples for a comprehensive understanding.

## Conclusion

Findings from this study have demonstrated that the psychometric properties of the Arabic WHO-5 in Lebanese adolescents are scientifically sound, supporting the usage of this easy-to-use, fast assessment tool in research. This scale is highly efficient for assessing wellbeing in adolescents. At the individual level, it would allow an easy, reliable, and valid evaluation of wellbeing, a core construct with strong correlations to improved mental and physical health, increased resilience, better academic performance, and stronger social relationships. On the social versant, it would be relevant to track overall levels of wellbeing in larger samples to prevent and reduce healthcare costs, enhance productivity, and foster a more stable thriving society, especially in times of political instability and ongoing turmoil.

## Data Availability

Data cannot be shared publicly. Data are available from the LAU Institutional Data Access / Ethics Committee for researchers who meet the criteria for access to confidential data.

## Declarations

### Human Ethics approval and Consent to participate declaration

The study was approved by the Institutional Review Board at the Lebanese American University, tracking number LAU.SAS.MJ1.28/Oct/2021, in accordance with the Declaration of Helsinki. Informed consent was obtained in writing by all participants and their parents before enrolling and filling the survey.

### Consent for publication

Not applicable

### Availability of data and materials

The datasets used and/or analyzes during the current study are available from the corresponding author on reasonable request.

### Authors’ contributions

M.SR, R.D, M.EK designed the study. R.D, M.EK wrote the main manuscript and S.H prepared the analyses and corresponding tables. All authors reviewed the manuscript.

## Acknowledgements

The authors wish to acknowledge all those who made the YEY project possible.

## REFERENCES

1. Keyes, C. L., Shmotkin, D., & Ryff, C. D. (2002). Optimizing well-being: the empirical encounter of two traditions. Journal of personality and social psychology, 82(6), 1007– 1022.

2. World Health Organization. Basic documents, 49th ed. Geneva: The Organization; (2020). p 1.

3. Bech, P., & Lundt, I. (2004). The WHO-5 reliability and validity in Danish general population surveys

4. Dupuy, H.J. (1977). The General Well-being Schedule. In I. McDowell & C. Newell (Eds.), Measuring health: a guide to rating scales and questionnaire (2nd ed): 206–213

5. Bonacchi, A., Chiesi, F., Lau, C., Marunic, G., Saklofske, D. H., Marra, F., & Miccinesi, G. (2021). Rapid and sound assessment of well-being within a multi-dimensional approach: The Well-being Numerical Rating Scales (WB-NRSs). PloS one, 16(6), e0252709. 10.1371/journal.pone.0252709

6. Ryff, C. D., & Keyes, C. L. B. (1995). The structure of psychological well-being in older adults: A longitudinal study. Journal of Personality and Social Psychology, 69(1), 140–154

7. Diener, E., Wirtz, D., Tov, W. et al. New Well-being Measures: Short Scales to Assess Flourishing and Positive and Negative Feelings. Soc Indic Res 97, 143–156 (2010). 10.1007/s11205-009-9493-y

8. Fekih-Romdhane, F., Cherif, W., Alhuwailah, A., Fawaz, M., Shuwiekh, H. A. M., Helmy, M., … & Hallit, S. (2023). Cross-Country Validation of the Arabic version of the WHO-5 Well-Being Index in non-clinical young adults from six Arab countries. 10.21203/rs.3.rs-2988215/v1

9. Perera, B. P. R., Jayasuriya, R., Caldera, A., & Wickremasinghe, A. R. (2020). Assessing mental well-being in a Sinhala speaking Sri Lankan population: validation of the WHO-5 well-being index. Health and quality of life outcomes, 18(1), 305. 10.1186/s12955-020-01532-8

10. Mendle, J., & Koch, M. K. (2019). The psychology of puberty: What Aren’t we studying that we should? Child Development Perspectives, 13(3), 166–172. 10.1111/cdep.12333

11. Heim, C., Newport, D.J. Mletzko, T. et al. (2008). The link between childhood trauma and depression: insights from HPA axis studies in humans. Psychoneuroendocrinology, 33: 693–710

12. Rapee, R.M., Oar, E.L., Johnco, C.J. et al. (2019). Adolescent development and risk for the onset of social-emotional disorders: a review and conceptual model. Behavior Research and Therapy, 123

13. Fenwick-Smith, A., Dahlberg, E.E. & Thompson, S.C. (2018). Systematic review of resilience-enhancing, universal, primary school-based mental health promotion programs. BMC Psychology 6, 30. 10.1186/s40359-018-0242-3

14. https://www.who.int/health-topics/adolescent-health#tab=tab_1

15. Ayyash-Abdo, H., & Sanchez-Ruiz, M. J. (2012). Subjective wellbeing and its relationship with academic achievement and multilinguality among Lebanese university students. International Journal of Psychology, 47(3), 192–202. 10.1080/00207594.2011.614616

16. Quansah, F., Hagan, J. E., Jr, Ankomah, F., Agormedah, E. K., Nugba, R. M., Srem-Sai, M., & Schack, T. (2022). Validation of the WHO-5 Well-Being Scale among Adolescents in Ghana: Evidence-Based Assessment of the Internal and External Structure of the Measure. Children (Basel, Switzerland), 9(7), 991. 10.3390/children9070991

17. McMahon, E. M., Corcoran, P., O’Regan, G., Keeley, H., Cannon, M., Carli, V., Wasserman, C., Hadlaczky, G., Sarchiapone, M., Apter, A., Balazs, J., Balint, M., Bobes, J., Brunner, R., Cozman, D., Haring, C., Iosue, M., Kaess, M., Kahn, J. P., Nemes, B., … Wasserman, D. (2017). Physical activity in European adolescents and associations with anxiety, depression and well-being. European child & adolescent psychiatry, 26(1), 111– 122. 10.1007/s00787-016-0875-9

18. Sirpal, M. K., Haugen, W., Sparle, K., & Haavet, O. R. (2016). Validation study of HSCL-10, HSCL-6, WHO-5 and 3-key questions in 14-16 year ethnic minority adolescents. BMC family practice, 17, 7. 10.1186/s12875-016-0405-3

19. Maalouf, F. T., Haidar, R., Mansour, F., Elbejjani, M., Khoury, J. E., Khoury, B., & Ghandour, L. A. (2022). Anxiety, depression and PTSD in children and adolescents following the Beirut port explosion. Journal of affective disorders, 302, 58–65. 10.1016/j.jad.2022.01.086

20. El Khoury-Malhame M., Bou Malhab S., Chaaya R., Sfeir M. and El Khoury S. (2024). Coping during socio-political Uncertainty. Frontiers in Psychiatry. 14: 126760

21. El Khoury-Malhame M., Rizk R, Joukayem E, Rechdan A, Sawma T. (2023). The psychological impact of COVID-19 in a socio-politically unstable environment: protective effects of sleep and gratitude in Lebanese adults. BMC Psychology;11(1):14. doi:10.1186/s40359-023-01042-4

22. El Hajj, S. (2020). Writing (from) the Rubble: Reflections on the August 4, 2020 Explosion in Beirut, Lebanon. Life Writing, 18(1), 7–23. 10.1080/14484528.2020.1830736

23. Sfeir, M., Akel, M., Hallit, S. et al. Factors associated with general well-being among Lebanese adults: The role of emotional intelligence, fear of COVID, healthy lifestyle, coping strategies (avoidance and approach). Curr Psychol 42, 17465–17474 (2023). 10.1007/s12144-021-02549-y

24. Sanchez-Ruiz, M. J., Tadros, N., Khalaf, T., Ego, V., Eisenbeck, N., Carreno, D. F., & Nassar, E. (2021). Trait Emotional Intelligence and wellbeing during the pandemic: The mediating role of Meaning-Centered Coping. Frontiers in Psychology, 12, Article 648401. 10.3389/fpsyg.2021.648401

25. Bhandari, N., & Gupta, S. (2024). Trends in Mental Wellbeing of US Children, 2019-2022: Erosion of Mental Health Continued in 2022. International journal of environmental research and public health, 21(2), 132. 10.3390/ijerph21020132

26. Johansen SK (1998): The use of well-beingmeasures in primary health care – the Dep-Care project; in World Health Organization,Regional Office for Europe: Well-Being Measures in Primary Health Care – the DepCareProject. Geneva, World Health Organization,1998, target 12, E60246.

27. Löwe, B., Decker, O., Müller, S., Brähler, E., Schellberg, D., Herzog, W., & Herzberg, P. Y. (2008). Validation and standardization of the Generalized Anxiety Disorder Screener (GAD-7) in the general population. Medical care, 46(3), 266–274. 10.1097/MLR.0b013e318160d093

28. Sawaya, H., Atoui, M., Hamadeh, A., Zeinoun, P., & Nahas, Z. (2016). Adaptation and initial validation of the Patient Health Questionnaire - 9 (PHQ-9) and the Generalized Anxiety Disorder - 7 Questionnaire (GAD-7) in an Arabic speaking Lebanese psychiatric outpatient sample. Psychiatry research, 239, 245–252. 10.1016/j.psychres.2016.03.030

29. Kroenke, K., Spitzer, R. L., & Williams, J. B. (2001). The PHQ-9: validity of a brief depression severity measure. Journal of general internal medicine, 16(9), 606–613. 10.1046/j.1525-1497.2001.016009606.x

30. Mundfrom DJ, Shaw DG, Ke TL (2005). Minimum sample size recommendations for conducting factor analyses. International journal of testing, 5(2):159–168.

31. Hu Lt, Bentler PM (1999). Cutoff criteria for fit indexes in covariance structure analysis: Conventional criteria versus new alternatives. Structural equation modeling: a multidisciplinary journal, 6(1):1–55.

32. Malhotra N, Dash S: Marketing Research: An Applied Orientation (2011). Pearson, Ed In.: Delhi

33. Chen FF: Sensitivity of goodness of fit indexes to lack of measurement invariance. Structural equation modeling: a multidisciplinary journal 2007, 14(3):464–504.

34. Vadenberg R, Lance C: A review and synthesis of the measurement in variance literature: Suggestions, practices, and recommendations for organizational research. Organ Res Methods 2000, 3:4–70.

35. Swami V, Todd J, Azzi V, Malaeb D, El Dine AS, Obeid S, Hallit S: Psychometric properties of an Arabic translation of the Functionality Appreciation Scale (FAS) in Lebanese adults. Body Image 2022, 42:361–369.

36. Dunn TJ, Baguley T, Brunsden V (2014). From alpha to omega: A practical solution to the pervasive problem of internal consistency estimation. British journal of psychology, 105(3):399–412.

37. Hair Jr JF, Sarstedt M, Ringle CM, Gudergan SP (2017). Advanced issues in partial least squares structural equation modeling: saGe publications.

38. El Khoury-Malhame M., Harajli, D., Rekowska, D., Jakubowska, M. and Ohm R. (2024) Can The Phoenix Still Rise? Traumatic Effect of Beirut Port Explosion on Lebanese People’s Experiences. Psychological Trauma: Theory, Research, Practice and Policy. Accepted

39. Loewenthal KM, Lewis CA (2018) An introduction to psychological tests and scales.Table 1. Categorization of participants according to their depression and anxiety scores. Psychology press.

40. Bech P (2004) Measuring the dimension of psychological general well-being by the WHO-5. Quality of life newsletter:15–16.

41. Awata, S., Bech, P. E. R., Yoshida, S., Hirai, M., Suzuki, S., Yamashita, M., … & Oka, Y. (2007). Reliability and validity of the Japanese version of the world health organization-five well-being index in the context of detecting depression in diabetic patients. Psychiatry and clinical neurosciences, 61(1), 112–119.

42. Ghazisaeedi, M., Mahmoodi, H., Arpaci, I., Mehrdar, S., & Barzegari, S. (2021). Validity, reliability, and optimal cut-off scores of the WHO-5, PHQ-9, and PHQ-2 to screen depression among university students in Iran. International Journal of Mental Health and Addiction, 1–10.

43. Krieger T., Zimmermann J., Huffziger S. Ubl B., Diener C.,, Kuehner C., Holtforth MG. (2014). Measuring depression with a well-being index: Further evidence for the validity of the WHO Well-Being Index (WHO-5) as a measure of the severity of depression. Journal of Affective Disorders,156: 240–244. 10.1016/j.jad.2013.12.015.

44. Topp, C. W., Østergaard, S. D., Søndergaard, S., & Bech, P. (2015). The WHO-5 Well-Being Index: a systematic review of the literature. Psychotherapy and psychosomatics, 84(3), 167–176. 10.1159/000376585

45. Middle East and North Africa. : Youth Facts. Available at: https://www.youth-policy.org/mappings/regionalyouthscenes/mena/facts/

